# Estimation of Survival with Parkinson’s disease in the Canadian Longitudinal Study on Aging: An analysis using only current disease durations observed at baseline

**DOI:** 10.1101/2025.06.30.25329557

**Authors:** James McVittie, David Wolfson, Ronald B. Postuma, Christina Wolfson

**Affiliations:** Department of Mathematics and Statistics, McGill University, Montreal, Quebec, Canada; Research Institute of the McGill University Health Centre, Montreal, Quebec, Canada; Department of Epidemiology, Biostatistics and Occupational Health, School of Population and Global Health, McGill University, Montreal, QC, Canada; Department of Medicine, McGill University, Montreal, QC, Canada; Department of Neurology and Neurosurgery, McGill University, Montreal, Quebec, Canada

**Keywords:** CLSA, Parkinson’s Disease, Survival, Current lifetimes, Stacking Model

## Abstract

Previous studies of survival with Parkinson’s disease (PD) have relied primarily on survival data from incident PD cohort studies with follow-up, prevalent PD cohort studies with follow-up, or a combination of these two study types. Each imposes logistical and resource constraints because of the length of follow-up required. Here, by using only the current disease durations for prevalent PD cases at study baseline we propose a strategy that does not require follow-up when estimating survival with PD. We apply our methods to data collected as part of the Canadian Longitudinal Study on Aging (CLSA). Using the reported disease durations for 110 CLSA participants classified as having prevalent Parkinson’s Disease at baseline, the estimated median survival was 5.72 years or 5.94 years, depending on how we accounted for possible uncertainty in the recalled diagnosis dates. Our results suggest a probability of at least 0.12 of surviving more than 15 years with PD. Our estimates are lower than those from other studies, which have entailed lengthy follow-up. We use only baseline data from a study that it is projected to last for twenty years. Under certain constraints our methods can be applied to other diseases and where “early” results are desired.

## INTRODUCTION

Parkinson’s disease (PD) is a debilitating neurodegenerative disorder affecting more than 6 million worldwide [1]. Researchers have reported reduced survival in individuals suffering from PD relative to the general population [2, 3] whether survival is estimated from perceived onset or from diagnosis. Differences in study design have resulted in a wide range of estimates [4].

The gold standard for estimating survival with PD requires the follow-up of an initially disease-free cohort to ascertain incident cases as they occur or are diagnosed and further follow-up of these cases to observe either death, loss to follow-up, or end of study. Consequently, incident cohort studies of survival with PD are resource- and time-intensive. An alternative to the incident cohort study with follow-up, is the so-called prevalent cohort study with follow-up. In such a study a cohort of individuals is first identified with prevalent PD (the prevalent cohort), and their ages of onset and/or diagnosis recorded; this cohort is then followed until death, loss to follow-up or end of study. There are two time-saving advantages to prevalent cohort studies with follow-up. First, the long follow-up time (to observe disease onset or diagnosis) of initially disease-free subjects in an incident cohort study is avoided. Second, in order to record full failure times, the shorter follow-up of the prevalent cases is only required; these are appended to the current disease durations that will have been ascertained at study baseline. In contrast, in an incident cohort study, follow-up is required from disease onset in order to ascertain full failure times. Nevertheless, follow up of the prevalent cases is still required and for diseases of relatively long duration it may be years before estimates of survival can be obtained. However, under the assumption that the incidence rate has remained relatively constant over time, it is possible to avoid the need for follow-up and estimate survival based only on the observed current disease durations in the prevalent cohort at study baseline [5, 6, 7]. While estimators based only on current disease duration are clearly not as efficient as those based on data that include prospective follow-up, there is a considerable benefit in being able to carry out statistical inference early on in a study. This is the case with the Canadian Longitudinal Study on Aging (CLSA). The CLSA is a prospective study of aging, launched in 2011 for which more than 50,000 Canadian residents aged 45-85 years of age were recruited to be followed for 20 years or until death or loss to follow up. In addition to a baseline assessment (2011-2015), participants are re-contacted every 3 years for repeat assessments. In the CLSA information was collected on a wide variety of variables including the presence of chronic diseases.

In particular, prevalent cases of PD were identified at baseline through a disease ascertainment algorithm that included a symptom screening questionnaire. With follow-ups of participants scheduled every 3 years, there is the potential to carry out an incident cohort study with follow-up of the participants who are PD-free at baseline. It will, however, take several follow-ups to accrue enough incident cases of PD and multiple follow-ups to be able to provide precise and unbiased estimates of survival. Therefore, while the follow-up of the prevalent cases is the ultimate goal this will mean a considerable delay before precise estimates of survival can be obtained. In this paper, while adjusting for length-bias, we propose a strategy for estimating survival with PD that does not require follow-up. We use only the reported current disease durations (more formally called, current lifetimes or backward recurrence times) of the prevalent cases as determined at CLSA baseline.

## METHODS

### The Canadian Longitudinal Study on Aging (CLSA)

The CLSA is a national study of adult development and aging, launched in 2011 with data collection waves every 3 years planned until 2033 [8]. At the study baseline (2011-2015), 51,338 participants aged 45-85 were recruited from all 10 Canadian provinces using 3 sampling frames: The Canadian Community Health Survey 4.2, provincial health registration data and random digit dialing. There are two modes and intensities of data collection in the CLSA: Participants in the Tracking Cohort (n=21,241) participate through 60-90-minute telephone interviews. Members of the Tracking cohort were recruited from all 10 Canadian provinces. The Comprehensive Cohort (30,097) participate through more detailed data collection that includes a 90 minute face-to-face in-home interview followed by a visit to one of 11 CLSA bespoke Data Collection Sites (DCS) for additional interviews, physical assessments, and blood and urine collection. These visits last 2-3 hours. The Data Collection Sites are located in 7 provinces. There are no DCS in Saskatchewan, New Brunswick or Prince Edward Island. At each wave of data collection, with few exceptions, the Tracking Cohort questionnaire and the questionnaires asked of members of the Comprehensive Cohort are identical in content, differing only in mode of administration. One such exception, relevant to this research, is described below.

### Identification of prevalent Parkinson’s disease cases

The CLSA questionnaires include single self-report questions for 23 chronic and non-neurological somatic or psychiatric conditions. Chronic conditions are defined as those that are ‘long-term’, i.e. those that are expected to last or have already lasted at least 6 months. The specific question asked at baseline for Parkinson’s disease was “Has a doctor ever told you that you had Parkinsonism or Parkinson’s Disease?”. This question was asked of all participants (n=51,338) at the baseline assessment. As a supplement to the single self-report question, a CLSA parkinsonism/PD questionnaire module was also applied to the 30,097 Comprehensive Cohort participants at the study baseline. This module was based on the screening questionnaire developed by Tanner [9, 10] and additional questions were asked concerning the use of medications typically prescribed for parkinsonism/PD. The Tanner questionnaire consists of 9 symptom questions seeking information about difficulty getting out of a chair, tremor, difficulty buttoning, smaller handwriting, softer voice, change in facial expression, lack of balance, “freezing”, and shuffling gait. A cutoff of 4 relative to the gold standard of an assessment by a movement disorder specialist has been found to have a sensitivity of 91% and a specificity of 81% for a diagnosis of parkinsonism [10]. The Tanner questionnaire is one of the most studied screening tools and has been translated into several languages [11]. In this study we classified individuals who (a) responded positively to the single question and scored more than 3 out of 9 on the screening questionnaire; or (b) responded positively to the single question and reported that they were taking medication for parkinsonism as having prevalent Parkinson’s disease at baseline (since PD medications can cause symptoms to resolve, pushing questionnaire scores below threshold). This algorithm has been used in other CLSA publications [7]. As part of the CLSA parkinsonism/PD algorithm participants who gave a “yes” answer to the self-report question were also asked “At what age, or in what year, did you first develop parkinsonism or were you first told you had Parkinson’s Disease?” This question left open the possibility that participants were being asked when they first experienced onset of parkinsonism. We have chosen, however, to interpret the ages provided by the participants as the recalled ages of diagnosis with PD, as this is far more likely to have been the age provided. This interpretation was based on two considerations: first, the answers were more likely to be associated with the preceding self-report question, which pertained to a prior *diagnosis* of parkinsonism or PD. Second, it is reasonable to assume that participants were more likely to recall their approximate age of diagnosis than their age of onset of a disease that often presents with insidious initial signs.

The CLSA parkinsonism/PD module was not included in the baseline interview for the Tracking participants and was only introduced into the Tracking questionnaire at the Maintaining Contact Interview, a brief interview that was conducted approximately 18 months after the baseline interview. Because the participants in the Tracking Cohort did not provide the information needed to complete the module at baseline as did the Comprehensive Cohort participants, only the CLSA Comprehensive Cohort participant data are used in our analysis.

## STATISTICAL METHODS

We began by assuming that the incidence rate of PD remained approximately constant over the roughly twenty years prior to 2011, as did the rate of diagnosis with PD. More precisely, the assumption is that the onsets of PD in the population from which the CLSA participants were drawn, follow a stationary Poisson process. In the survival analysis literature this is known as the *stationarity assumption* [12]. This, or that the incidence rate is non-constant but known, is needed in order to estimate the failure time distribution using only the current disease durations of participants at baseline [13]. The stationarity assumption was deemed reasonable because the literature is inconsistent as to whether the incidence rate has increased, decreased, or remained constant [11, 14, 15, 16, 17]. Further, it is not possible to carry out a test for stationarity using only the current disease durations.

In the statistical methodology literature, estimation using the current disease durations has received considerable attention and is divided into two broad categories: the failure time distribution is assumed to be non-parametric [5, 6, 7], or is parametrically defined [5, 6]. Because non-parametric estimators tend to suffer from instability and sometimes give very wide confidence intervals [5, 6] we chose a parametric approach with two modifications of the basic methods described in the literature.

The data collected at baseline as part of the CLSA were assumed to comprise the recalled diagnosis dates for those participants who were classified as having prevalent PD (see *Identification of prevalent Parkinson’s disease cases)*. Therefore, it was possible to compute “raw” current disease durations from diagnosis, which by definition, were uncensored. However, we did not use these data directly to estimate the failure time distribution. Rather, we adjusted them in anticipation of possible uncertainty in the recalled dates of diagnosis. We reasoned that the further the reported diagnosis date was from the baseline date the more likely the recalled date would be in error as would the true date of diagnosis. We supposed that since the onset of PD is insidious true diagnosis was likely to have occurred at an earlier time than that recalled. For comparison we allowed respectively for no lapses in recall, a small lapse in recall, and a larger lapse in recall. Specifically, if a participant’s computed current disease duration was t, this was first taken as the true current disease duration, for the estimation of survival. Second, we increased t by an amount drawn from an exponential distribution with mean t/4 before estimating survival. Third, we increased t by an amount drawn from an exponential distribution with mean t/2. Thus, the adjustment was made under the assumption that the shorter t, the more trust one would have in the recall and the smaller the adjustment needed.

We used two separate parametric models for the failure time distribution initially, the Weibull and the Gamma since they are by far the most commonly used survival distributions that allow for an increasing hazard. With expression 4 in [18, pg.4] as the likelihood function we estimated the failure time distributions for each parametric model, by finding the maximum likelihood estimates of the respective parameters using the amended data as described above. Because we were concerned about model misspecification and the consequent possible non-robustness of any single parametric model, we used a stacking procedure [19] to construct a model that included both of the above parametric models, weighted using the Brier score [18]. By implementing parametric bootstraps, we found pointwise 95% confidence intervals for the failure time distributions using the separate parametric models and the stacked model.

## RESULTS

A total of 111 participants in the Comprehensive Cohort were identified as having prevalent Parkinson’s disease at the CLSA baseline. The flow chart (Figure 1) shows the paths, described in the Methods section, leading to inclusion of CLSA participants in the Prevalent Parkinson’s disease cohort. For one participant the computed current disease duration was negative, and this participant was excluded, leaving 110 for analyses. Of these 110 subjects, 6 reported durations of 0, no doubt corresponding to a diagnosis that had occurred less than a year prior to the baseline interview. We imputed strictly positive durations for these subjects by taking independent draws from a Uniform distribution on the interval (0, 1). Below, Tables 2 and 3, and Figures 2,3, and 4 refer to the Stacked model.

**Figure 1:**
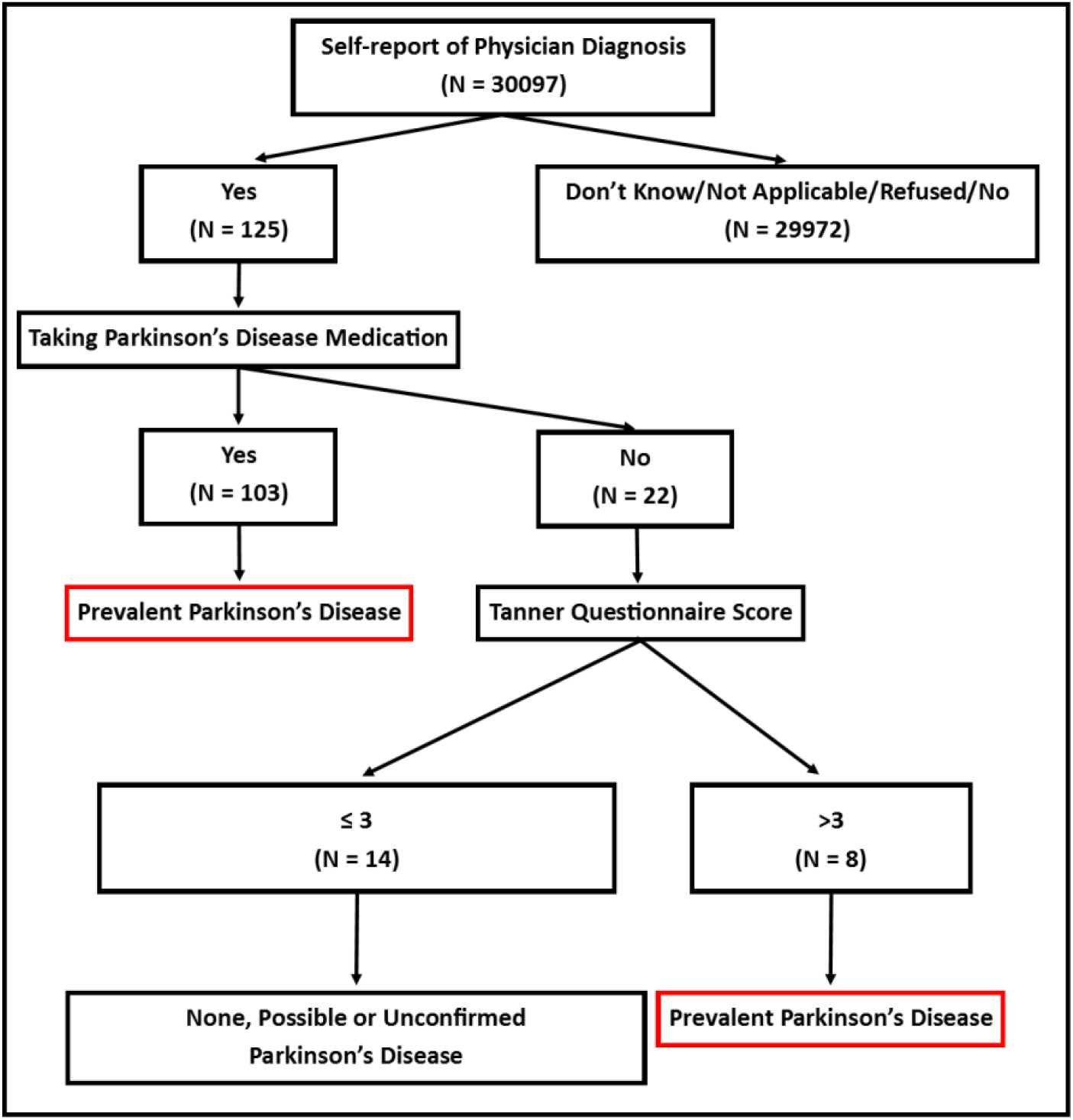
A flowchart of the identification of participants with Prevalent Parkinson’s Disease in the CLSA Comprehensive Cohort.

**Figure 2:**
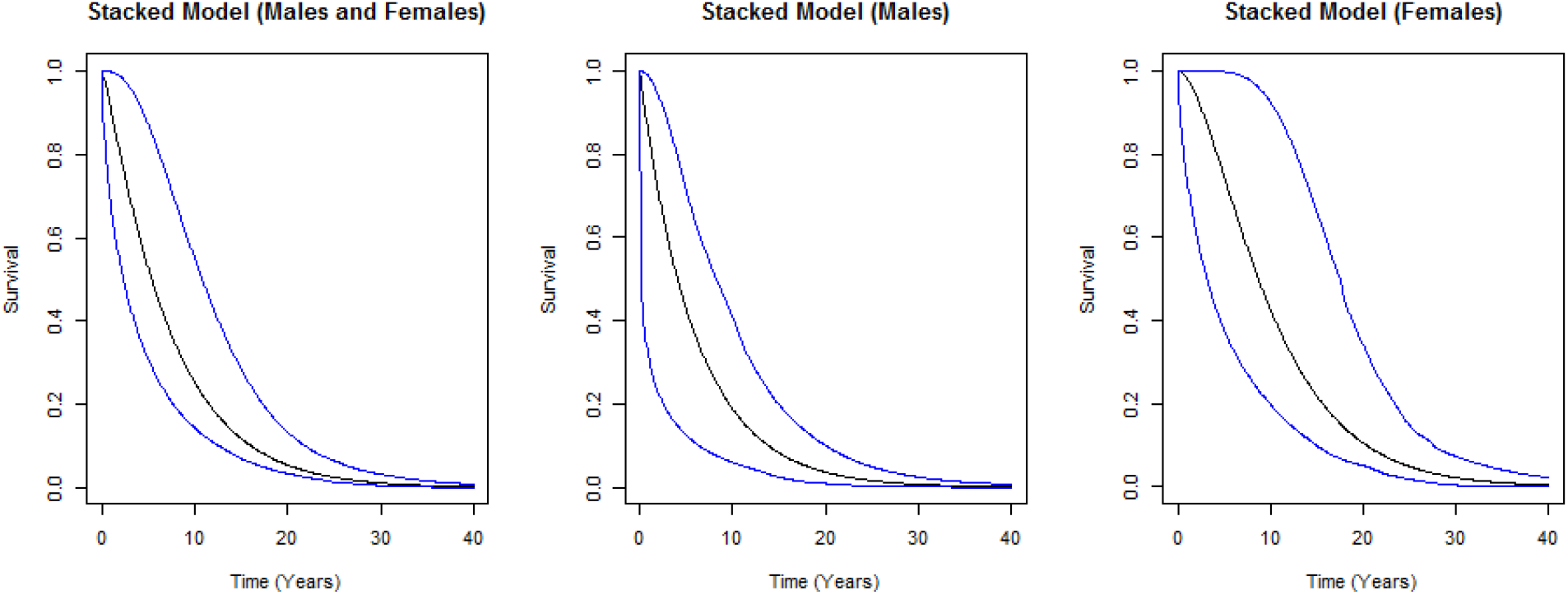
The estimated failure time distribution and pointwise 95% confidence intervals (outer bands) based on the Stacked model stratified by sex with no adjustment. The stack includes the Weibull and Gamma distributions with respective Stacked model weights of (9.63×10^−7^, 0.999) for the cohort of males and females combined, (9.73×10^−8^, 0.999) for males and (6.97×10^−6^, 0.999) for females

**Figure 3:**
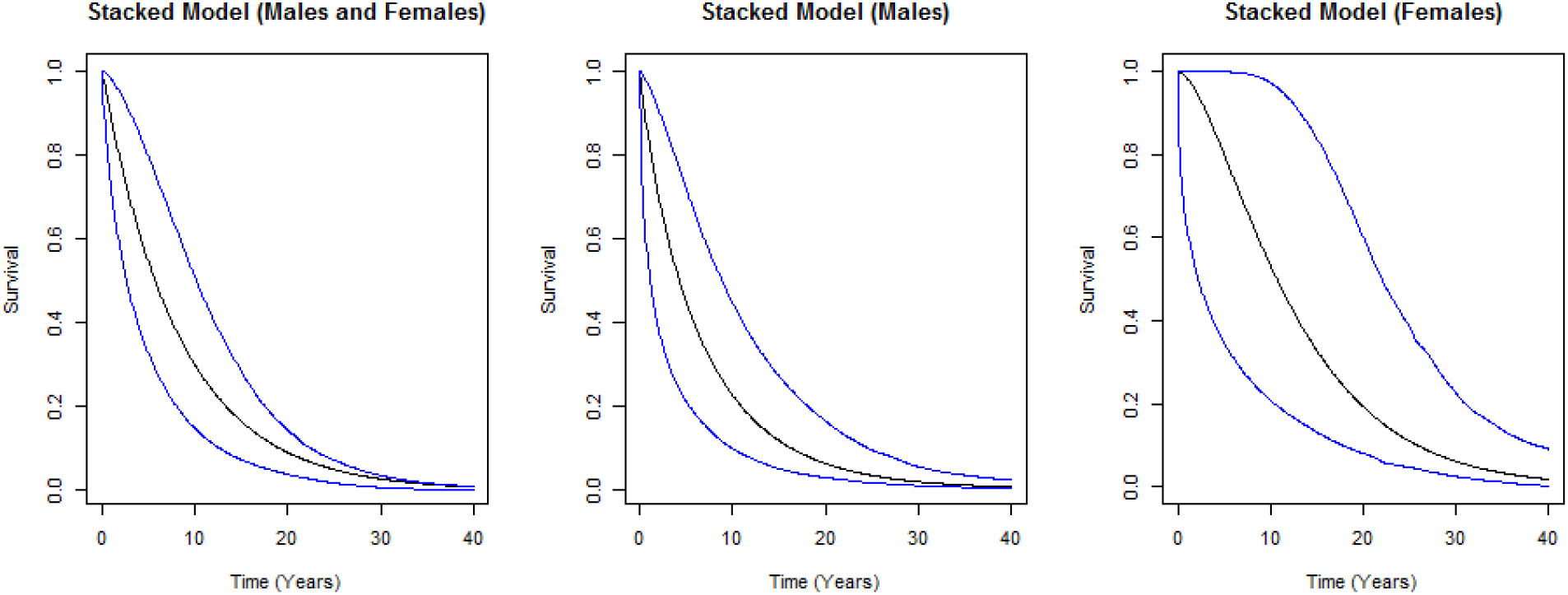
The estimated failure time distribution and pointwise 95% confidence intervals (outer bands) based on the Stacked model stratified by sex including Exponential adjustment with mean t/4. The stack includes the Weibull and Gamma distributions with respective Stacked model weights of (0.75, 0.25) for the cohort of males and females combined, (0.89, 0.11) for males and (0.07, 0.93) for females

**Figure 4:**
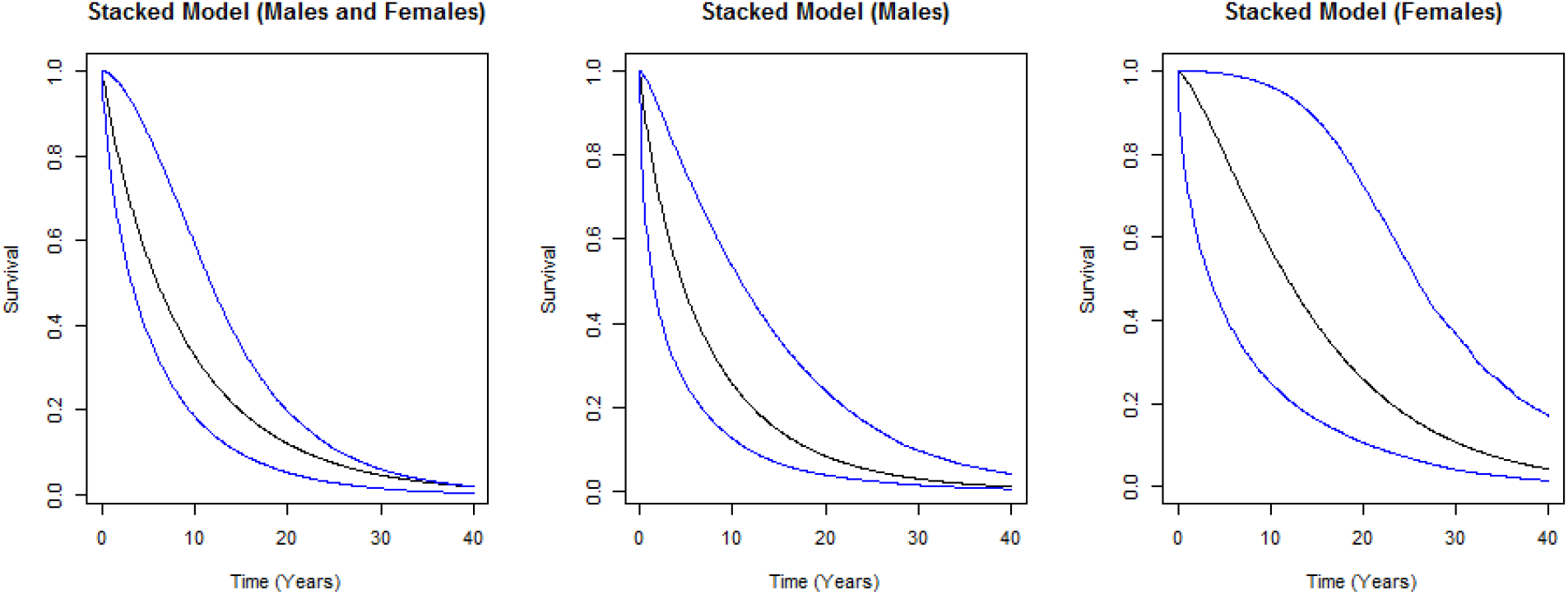
The estimated failure time distribution and pointwise 95% confidence intervals (outer bands) based on the Stacked model stratified by sex including Exponential adjustment with mean t/2. The stack includes the Weibull and Gamma distributions with respective Stacked model weights of (0.92, 0.08) for the cohort of males and females combined, (0.97, 0.03) for males and (0.32, 0.68) for females

Table 1 shows that the respective reported quartiles for the ages at diagnosis were similar for males, females, and males and females combined. The median ages of those identified with PD at CLSA baseline were identical (71 years) for males and females. Females reported a slightly younger median age at onset/diagnosis than males (62 years compared to 64.5 years respectively).

**Table 1:** Summary statistics (first quartile (Q1), median (Q2), third quartile (Q3)) of age at onset/diagnosis and age at CLSA baseline for subjects classified as having Prevalent Parkinson’s Disease.

| Variable | Males (N=70) | Females (N=40) | Total (N=110) |
| --- | --- | --- | --- |
| Age at onset/diagnosis (Q1, median, Q3) | 56.25, 64.50, 72.00 | 55.50, 62.00, 69.25 | 56.00, 63.00, 71.00 |
| Age at CLSA baseline (Q1, median, Q3) | 63.25, 71.00, 77.00 | 63.00, 71.00, 76.25 | 63.00, 71.00, 77.00 |

The estimated median survival times with PD (and confidence intervals) for each of the adjustment categories are presented in Table 2. Overall (for Males and Females combined) the estimated median survival ranged from 5.34 and 5.94 years. For Males the range was 4.07 years to 4.52 years and for Females 8.07 years to 11.94 years. We elaborate on the main features of our results under the heading “Discussion.”

**Table 2:** Estimated median survival with 95% confidence intervals using the Stacked model with no adjustment; exponential adjustment with mean t/4; exponential adjustment with mean t/2.

| Adjustment | Median | 95% Confidence Interval |
| --- | --- | --- |
| *No adjustment* |  |  |
| Males | 4.07 | (0.1, 8.2) |
| Females | 8.07 | (3.2, 17.5) |
| Overall | 5.34 | (2.3, 10.8) |
| *t/4 adjustment* |  |  |
| Males | 4.25 | (1.1, 8.8) |
| Females | 10.69 | (2.2, 22.0) |
| Overall | 5.72 | (2.6, 10.2) |
| *t/2 adjustment* |  |  |
| Males | 4.52 | (1.4, 10.9) |
| Females | 11.84 | (3.5, 25.7) |
| Overall | 5.94 | (3.2, 11.7) |

Table 3 displays the estimated 5-year, 7-year, 12-year and 15-year survival probabilities, and 95% pointwise confidence intervals. These results are presented for the model with no adjustment, mean t/4 adjustment and mean t/2 adjustment. This table along with accompanying Figures 2, 3, and 4 are our primary results. Figures 2, 3 and 4 display, respectively, the estimated failure time distributions, and bootstrapped pointwise 95% confidence intervals for the Stacked model for the three adjustment scenarios. Each figure includes the distributions for males and females combined and separately. The Stacked model includes the Weibull and Gamma distributions in the stack and the weights given to each model in the stack are provided in the legends to these figures. We found that females had longer survival than males in all adjustment scenarios. Our results suggest a probability of 0.12 of surviving more than 15 years with no adjustment and 0.16 and 0.20 with adjustments t/4 and t/2 respectively. We also observed important differences in the survival probabilities between males and females with the survival probabilities for the selected time points higher for females than males.

**Table 3:**
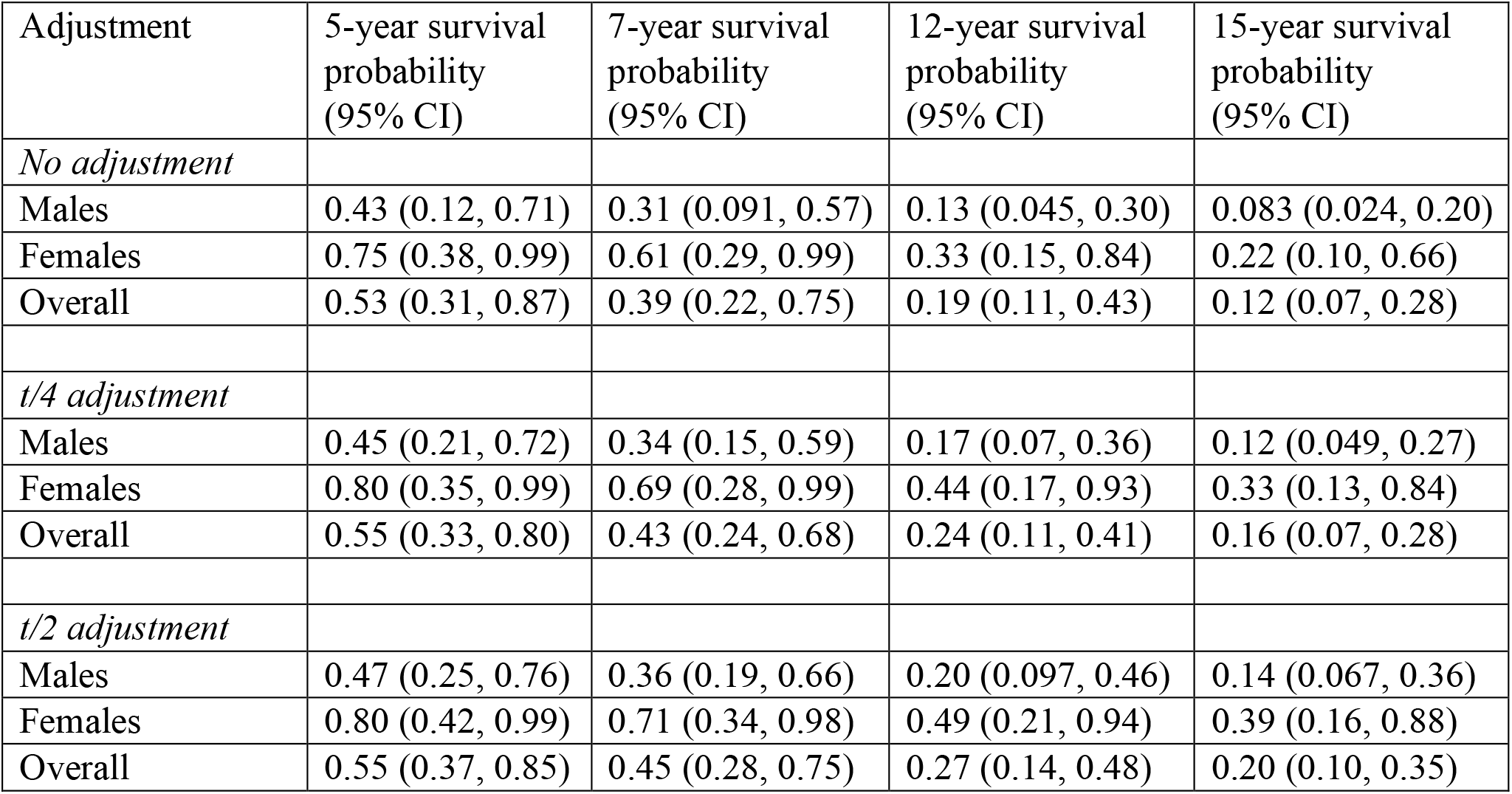
Estimated survival probabilities with 95% pointwise confidence intervals using the Stacked model with no adjustment; exponential adjustment with mean t/4; exponential adjustment with mean t/2

## DISCUSSION

For males and females combined our range of estimated median survival times from diagnosis with PD, is lower than most estimates in the literature. (See [4] for an extensive systematic review of the literature on survival with PD). While we also found shorter survival in males than females as have others [3, 20], in our work the median survival for females was consistently twice that of males. Our concern was that the magnitude of the difference is anomalous. For this reason, we searched for an explanation, beginning with an inspection of the data, examining the recorded current disease durations. We found that a far higher proportion of males than females reported current disease durations of either 0, 1 or 2 years; thereafter the reported current disease durations were similar. We then checked whether the actual calculations of the current disease durations from the raw data provided by the participants were correct: we found no errors. We also stratified the current disease durations by province, to assess whether the anomalous data may have come from one or two CLSA data collection sites perhaps reflecting a systematic error in data collection; this was not the case. Finally, as a very rough check, we computed the current disease durations from the Tracking cohort data, even though for reasons described in the Methods section, these data were not complete enough for our analysis; the current disease durations from the Tracking cohort were consistent with those from the Comprehensive cohort. Therefore, our only explanation is that our data are simply anomalous.

In summary, using data from the CLSA Comprehensive cohort, and applying a Stacking model with adjustment (t/4 and t/2) for uncertainty in the reported diagnosis/onset times we estimated a range of median survival from 5.72 to 5.94 years from Parkinson’s disease diagnosis using only current disease duration at CLSA baseline. The estimates are slightly shorter than those reported in the literature, although there is a wide range of estimated median survival in studies with Parkinson’s disease (See [4] for an extensive systematic review of the literature on survival with PD).

In estimating the 5-, 7-and 12-year survival probabilities we noted wide 95% pointwise confidence intervals which could be due to at least three reasons. First, the course of Parkinson’s disease is known to vary considerably between subjects. Second, the sample sizes used in the analysis were rather small (only 70 males and 40 females). Third, our inference is based on the current disease duration, the only information that is available at the study baseline. The resulting “missingness” of the remaining disease duration data, given that there had not yet been follow-up, adds uncertainty to the statistical inference.

We are able to estimate the failure time distributions in the CLSA using only current disease durations from diagnosis because we have assumed that the onset process is a stationary Poisson process (that is, that PD has a constant incidence rate) [14, 16, 17]. However, the assumption of stationarity is not essential, as is pointed out by Lund [13]. If the onset process is a non-stationary Poisson process, provided the incidence rate is known, by a transformation of the time scale it is possible to reduce the non-stationary case to the stationary case [13]. Nevertheless, we chose not to allow for non-stationarity because of the disparity in the incidence rates of Parkinson’s disease over time reported in the literature, including some research that asserts a constant incidence rate overall [17] or in women [14, 16]. For the same reason we did not carry out simulations to examine the effect of assuming stationarity, when in fact the onset process is non-stationary.

Our study has some limitations that might have influenced our findings. The observation of lower estimated median survival than in other studies could be due to differences in the target populations sampled, to the statistical methods used to estimate survival, and to the definition of Parkinson’s disease, for example. In particular, it should be noted that two of the CLSA exclusion criteria at baseline (living in an institution and free from cognitive impairment) would likely have resulted in individuals at the latter stages of PD (i.e., with dementia and/or institutionalized) and thus possibly *longer* disease duration being excluded from the study. Once declared eligible for the CLSA, individuals had to agree to participate and successful enrolment into the Comprehensive cohort required not only an in person visit at home but also a visit to the CLSA Data Collection Site in their location. Thus, at study recruitment there is a possibility that individuals with PD/parkinsonism suffering from mobility problems might be less likely to agree to participate and/or travel to the Data Collection Site. These two features (CLSA eligibility criteria and physical requirements to participate) may have resulted in an excess of participants with *short* current disease durations. This selection bias would lead to reduced estimates of survival.

We must also consider that while we used the information collected in a PD/parkinsonism module to classify individuals with Prevalent Parkinson’s Disease, this strategy may have failed to pick up participants with PD or perhaps more importantly led us to include individuals in our sample of 110 who should not have been included and who may have overrepresented either short survival or long survival. Without a full clinical assessment this will always be a concern in observational studies that do not include diagnostic assessments for chronic conditions [21]. This concern has been studied by others and Tanner et al [22], in a case control study to examine the role of pesticides in the etiology of PD and parkinsonism, compared a simple self-report of PD with clinical assessment and found that 84% of those who self-reported PD were verified via clinical assessment and 100% of those selected who did not report PD, were found to be PD free. In our study we supplemented the self-report by requiring that participants were also taking medication for PD/parkinsonism or scored >3 on a parkinsonism screening tool.

The main contribution of the methods presented here is that they allow inference about survival with Parkinson’s disease in a cohort study from data collected at baseline, before follow-up has started. Our methods, however, are not restricted to survival with Parkinson’s disease and may be applied when early results are desired at the start of longitudinal studies. This could be particularly valuable when follow-up over a long time period is anticipated, or even in cross-sectional studies without follow-up.

## Data Availability

Data are available from the Canadian Longitudinal Study on Aging (www.clsa-elcv.ca) for researchers who meet the criteria for access to de-identified CLSA data.

## Acknowledgements

This research was made possible using the data collected by the Canadian Longitudinal Study on Aging (CLSA). Funding for the Canadian Longitudinal Study on Aging (CLSA) is provided by the Government of Canada through the Canadian Institutes of Health Research (CIHR) under grant reference: LSA 94473 and the Canada Foundation for Innovation, as well as the following provinces, Newfoundland, Nova Scotia, Quebec, Ontario, Manitoba, Alberta, and British Columbia. This research has been conducted using the CLSA Baseline Tracking Dataset version 3.5, Baseline Comprehensive Dataset version 4.1. Follow-up 1 Tracking Dataset version 2.1, and Follow-up 1 Comprehensive Dataset version 3.0, under Application Number 1909037. The CLSA is led by Drs. Parminder Raina, Christina Wolfson and Susan Kirkland. The analyses conducted here were funded by a CIHR Catalyst Grant awarded to C. Wolfson. JM was supported by the Natural Sciences Engineering Research Council of Canada (PGSD-3). The authors gratefully acknowledge the time and commitment of the CLSA participants, without whom this research would not be possible.

## Disclaimer

The opinions expressed in this manuscript are the author’s own and do not reflect the views of the Canadian Longitudinal Study on Aging.

